# Opportunities for targeted HIV prevention programs: measuring prevention gaps at public health facilities and social venues in Malawi

**DOI:** 10.64898/2026.08.19.26360772

**Authors:** Confidence Banda, Sarah Bourdin, Emmanuel Singogo, Evaristar Kudowa, Maganizo Chagomerana, John Chapola, Harriet Jones, Thomas Hartney, Jessie K. Edwards, Andreas Jahn, Gift Kawalazira, Yohane Kamgwira, Lucy Platt, Brian Rice, James R. Hargreaves, Mina C. Hosseinipour, Sharon S. Weir

## Abstract

Precision targeting is essential for maximising impact and cost-effectiveness of interventions at this stage of the HIV response in Malawi. We aimed to measure gaps in access to and use of condoms, HIV testing, pre-exposure prophylaxis (PrEP) and voluntary medical male circumcision among HIV-negative individuals at public health facilities and social venues (bars, rest houses and liquor stores) in Blantyre, Malawi. We analysed cross-sectional data from 2,227 HIV-negative patients at government clinics and 1,634 patrons at social venues recruited in the Clinic vs Venue (CLOVE) study between January and March 2022. We estimated gaps in access to and use of condoms, HIV testing, PrEP and circumcision. Estimates were stratified by risk group, defined as reporting transactional sex, having multiple sex partners in the past 4 weeks, being female aged 15 to 24, or being male aged 30 and above. Access and use were based on self-reports. Overall, 30% of clinic and 60% of venue participants reported higher risk. Among men, we found a gap between access to condoms and condom use at last sex (76.7% vs 29.8% among clinic men; 75.4% vs 36.7% among venue men). Among women, the gap between access and use of condoms was 65.9% vs 18.0% at clinics and 79.9% vs 46.0% in the venues. Approximately 80-85% of participants reported knowing where to get an HIV test in Blantyre but less than half reported testing in the past 6 months. Use of PrEP was low (∼2%). Comparable proportions of men who paid for sex and those with multiple partners (∼77%) reported being circumcised, but this was lower among those aged 30 years or older (∼57%). Despite expanded HIV prevention services in Blantyre, gaps remain in the uptake of prevention services among people reachable at public health facilities and social venues. Use of PrEP was particularly low across all groups. Condom and testing use remained suboptimal despite high reported access. Targeted efforts are needed to address barriers to uptake, particularly for PrEP among high-risk venue-based populations.

## Introduction

Despite achievements in scaling up access to HIV treatment and recent declines in new HIV infections, globally HIV remains a public health concern, with an estimated 1.3 million new HIV infections in 2022 [1]. An estimated 14,000 new HIV infections occurred in Malawi in 2023, with an approximate annual incidence of 1.2 per 1,000 among adults (15-49 years) [1,2]. HIV incidence has declined steeply since 2010 in all districts across Malawi. Incidence is closely correlated with population density and the two districts with the main urban centres (Lilongwe and Blantyre) were estimated to account for 2,000 and 1,900 new infections in 2023 [3]. In the year of the study (2022), there was a reported annual incidence of 2.9 and 1.6 per 1,000 women and men aged 15-49 years, respectively in Blantyre district [3].

UNAIDS recommends prioritizing HIV prevention and allocating 25% of global HIV spending towards primary prevention interventions [4]. Access to prevention services remains uneven [5] Stigma, punitive laws, violence and discrimination present barriers to essential prevention services for marginalised populations including adolescent girls and young women, women engaging in sex work and transgender populations [4]. Consequently, the HIV pandemic continues to affect people in these populations more than general populations[6]. It is essential to understand the need for prevention services and to address access barriers to effectively reduce HIV transmission [7–10]. A systematic review of the effectiveness of HIV prevention interventions showed strong evidence for the efficacy of pre-exposure prophylaxis (PrEP) and voluntary medical male circumcision in primary studies of direct prevention mechanisms, but much less well-developed evidence of how to maximise the population impact of these tools [11].

Significant investments in HIV prevention have been made over many years in Blantyre, but there is an indication that considerable access and coverage gaps remain [12]. The Priorities for Local AIDS Control Efforts (PLACE) is an assessment tool to monitor and improve AIDS prevention program coverage in areas where HIV transmission is most likely to occur and has been deployed in Malawi to identify high-risk venues in 20 districts since 2016 [13]. Consistent with findings from other countries, critical members of sexual transmission networks, such as mobile, stigmatized, and hard-to-reach populations, have been identified at these venues [13–16]. While PLACE studies have historically focused on social venues, less is known about how prevention gaps at venues compare to those at health facilities. Clinics serve large volumes of individuals, including adolescent girls and young women (AGYW), who may also have unmet prevention needs. Comparing the two settings can help programmes determine where to allocate prevention resources for greatest impact.

The HIV prevention cascade is a useful framework to define the number of people in need and those who are receiving prevention services. It can further be used to determine who needs an HIV prevention service but cannot access it, and who does not use the service effectively. Tailoring the design of interventions to address these gaps can strengthen the impact of prevention interventions [15,17]. In this study we aimed to identify access and use gaps for condoms, HIV testing, PrEP and circumcision among higher-risk groups at both clinics and venues. We compared prevention gaps across settings to inform programme targeting decisions.

## Methods Study

### design

We analysed cross-sectional data collected as part of the Clinic vs Venue Testing: An Assessment of Expanded HIV Testing and Surveillance to Improve HIV Prevention in Blantyre (CLOVE) study, described in detail elsewhere (supplemental information) [18]. We accessed the study data on 7^th^ January 2024 which was anonymized before being accessed. In brief, data were collected in Blantyre district from January to March 2022 from patients at government clinics and patrons and workers at social venues. Venues were identified through a mapping exercise involving over 400 community informants applying the PLACE methodology [18]. At each selected venue, all consenting patrons present during predefined survey periods were invited to participate. At clinics, patients presenting for services during scheduled survey sessions were recruited consecutively. Interviewer-administered bio-behavioural surveys were conducted among venue patrons (n=1,802) and clinic patients (n=2,313).

### Study population

This analysis is limited to CLOVE study participants (n=3,861) with a non-reactive HIV test (Alere Determine^TM^). Participants were categorized into one or more risk categories, reflecting priority groups identified by the Malawi National Strategic Plan for HIV and AIDS 2020-2025 [19]:

1. Female sex workers (FSW) defined as women who answered affirmatively to: “Sometimes women have sex with someone in exchange for gifts or help with expenses or cash money. Think about the past 12 months. In the past 12 months, have you received gifts or favors or cash money in exchange for sex?”
2. Male clients of sex workers defined as men who answered affirmatively to: “In the past 12 months have you paid a woman to have sex with you?”
3. Women with multiple partners defined as those who reported more than one partner when asked how many men they had sex with
4. Men with multiple partners defined as men who reported more than one partner when asked how many women they had sex with in the past four weeks
5. Adolescent girls and young women (AGYW) defined as females aged 15-24 years.
6. Older men defined as males aged 30 years and above.

### Prevention interventions

We defined prevention interventions for the six risk categories described above as well as all male and female attenders of venues and clinics. Definitions of need, access and use for each prevention service and the reference populations are shown in Table 1. Guided by national policy, populations needing PrEP included: men who paid for sex, female sex workers, STI clinic patients, and AGYW reporting multiple sex partners. STI clinic patients were those who reported affirmatively to: “In the past 12 months, did a medical provider test or examine you to see if you had a sexually transmitted infection?”. We asked respondents whether they were taking PrEP but not about access to PrEP as it was in its initial phase of roll-out.

**Table 1.** Population groups for each prevention service and definitions of need, access and use for each cascade.

|  | **Clinic & Venue Risk Categories** | **Definition of Need** | **Definition of Access** | **Definition of Use** |
| --- | --- | --- | --- | --- |
| **Condoms** | • All women<br>• FSW<br>• Women with multiple partners<br>• AGYW<br>• All Men<br>• Clients of FSW<br>• Men with multiple partners<br>• Men aged 30+ | Sexually active defined as reporting vaginal or anal sex in the last 12 months. | Defined as answering “Easy” to the question: “If you wanted a condom, would it be easy or difficult for you to get one quickly?” | *Ever used* defined as answering “Yes” when asked: “Have you ever used a condom during vaginal sex?”<br>*Used last time* defined as answering “Yes” when asked: “The last time you had vaginal sex, did you use a condom?”<br>*Consistent use* defined as selecting: “I used condoms every time I had vaginal sex.” When asked: “Which best describes your condom use during vaginal sex in the past 6 months?” |
| **HIV Testing** | • All women<br>• FSW<br>• Women with multiple partners<br>• AGYW<br>• All Men<br>• Clients of FSW<br>• Men with multiple partners<br>• Men aged 30+ | Sexually active defined as reporting vaginal or anal sex in the last 12 months. | Defined as answering “Yes” to the question: “Do you know where to go to get tested for HIV in Blantyre?” | Defined as answering “Yes” when asked: “In the past 6 months, have you been tested for HIV and received your test results?” |
| **PrEP** | • FSW<br>• Clients of FSW<br>• STI Patients<br>• AGYW | Priority groups for PrEP according to national PrEP guidelines[20]:<br>• FSW<br>• Clients of FSW<br>• STI Patient<br>• AGYW who reports more | Not asked. | Defined as answering “Yes” to the question: “Are you currently taking PrEP to prevent getting an infection?” |

|  | Clinic & Venue Risk Categories | Definition of Need | Definition of Access | Definition of Use |
| --- | --- | --- | --- | --- |
|  |  | than one sexual partner |  |  |
| **VMMC** | • All Men<br>• Clients of FSW<br>• Men with multiple partners<br>• Men Age 30+ | All men | Not asked. | Defined as answering Yes to the question: “Are you a circumcised man?” |

### Analysis

We firstly described the demographic and sexual behaviour characteristics, stratified by recruitment site (venue vs. clinic) and gender (men vs women). Secondly, we summarised need, access and use of the prevention cascades for each risk group stratified by recruitment site and gender. Table 1 summarises the definitions used for need, access and use, with all those meeting the definition of having need as the overarching denominator. In addition, we separately estimated the percentage of respondents who needed and used the service but didn’t report accessing it. We created visual prevention cascades for each risk group stratified by recruitment site.

The study used a two-stage time-location sampling (TLS) design, consistent with standard practice in PLACE assessments and bio-behavioural surveys among populations at higher risk of HIV [21]. In the first stage, venues and clinics were sampled from a comprehensive frame constructed through the PLACE mapping exercise. In the second stage, individuals present at selected sites during defined survey periods were recruited. Survey weights were calculated as the inverse of the joint probability of selection at each stage. The first component was the probability that the venue or clinic was sampled. The second was the probability that the individual was sampled from within that site at the time of the survey. Weighted percentages and 95% confidence intervals are reported throughout. All counts are unweighted.

### Ethics

Participants consented before enrolment into the study and were given the choice to refuse any question or stop at any time. We obtained ethics approval from the National Health Sciences Research Council in Malawi (Protocol #:20/12/2637), the University of North Carolina in Chapel Hill, North Carolina (Protocol #: 22018), and the London School of Hygiene and Tropical Medicine (Ethics Ref #22959).

## Results

Among the 3,861 HIV negative participants included in this analysis, 57.7% were from clinics (n=2,227) and 42.3% were from venues (n=1,634) as seen in Table 2. Clinic and venue-based samples differed in the following key ways: proportionally more clinic patients were female (74.4% vs 24.5%); aged 15-24 (47.6% vs 35.9%); married (60.3% vs 41.0%); and not employed (68.9% vs 49.6%) than venue attenders. Over half of participants at clinic and venues lived in urban Blantyre.

**Table 2.** Descriptive and sexual behaviour characteristics of Men and Women at Clinics and Venues.

|  | Clinic Men |  |  | Clinic Women |  |  | Venue Men |  |  | Venue Women |  |  |
| --- | --- | --- | --- | --- | --- | --- | --- | --- | --- | --- | --- | --- |
|  | N | % | 95% CI | N | % | 95% CI | N | % | 95% CI | N | % | 95% CI |
| Total | 725 | 100.0 |  | 1502 | 100.0 |  | 1326 | 100.0 |  | 308 | 100.0 |  |
| Age group |  |  |  |  |  |  |  |  |  |  |  |  |
| 15-24 yrs. | 299 | 42.8 | 34.7 ; 50.9 | 709 | 49.3 | 43.1 ; 55.5 | 439 | 34.2 | 29.8 ; 38.7 | 140 | 40.9 | 29.0 ; 52.8 |
| 25-29 yrs. | 144 | 18.9 | 15.6 ; 22.3 | 316 | 20.9 | 18.8 ; 22.9 | 328 | 24.5 | 21.7 ; 27.4 | 69 | 21.1 | 12.9 ; 29.3 |
| ≥30 yrs. | 282 | 38.3 | 29.9 ; 46.7 | 477 | 29.8 | 23.6 ; 36.1 | 559 | 41.2 | 36.6 ; 45.8 | 99 | 38.0 | 27.7 ; 48.3 |
| Missing | - | - | - | - | - | - | - | - | - | - | - | - |
| Marital Status |  |  |  |  |  |  |  |  |  |  |  |  |
| Married or<br>Living with Partner | 336 | 46.3 | 36.4 ; 56.2 | 986 | 65.2 | 58.9 ; 71.4 | 617 | 45.1 | 40.9 ; 49.4 | 91 | 28.4 | 19.2 ; 37.6 |
| Separated | 24 | 3.9 | 2.1 ; 5.7 | 68 | 4.8 | 3.3 ; 6.3 | 72 | 5.3 | 3.9 ; 6.7 | 33 | 11.9 | 4.7 ; 19.2 |
| Divorced | 43 | 4.9 | 2.9 ; 6.9 | 150 | 9.2 | 6.9 ; 11.5 | 141 | 10.0 | 8.0 ; 12.0 | 85 | 24.6 | 16.2 ; 33.0 |
| Widowed | 2 | 0.3 | 0.0 ; 0.7 | 35 | 2.1 | 0.8 ; 3.5 | 5 | 0.5 | 0.0 ; 1.0 | 7 | 4.0 | 0.8 ; 7.3 |
| Never Married or<br>lived with a partner | 318 | 44.1 | 33.0 ; 55.2 | 260 | 18.5 | 12.0 ; 25.0 | 488 | 38.8 | 34.3 ; 43.3 | 88 | 29.6 | 21.4 ; 37.7 |
| Refused | 2 | 0.5 | 0.0 ; 1.5 | 3 | 0.2 | 0.0 ; 0.5 | 1 | 0.0 | 0.0 ; 0.1 | 2 | 0.7 | 0.0 ; 1.7 |
| Don't Know | - | - | - | - | - | - | 2 | 0.2 | 0.0 ; 0.6 | 2 | 0.8 | 0.0 ; 2.4 |
| Missing | 2 | 0.5 | 0.0 ; 1.5 | 3 | 0.2 | 0.0 ; 0.5 | - | - | - | - | - | - |
| Completed Secondary<br>School |  |  |  |  |  |  |  |  |  |  |  |  |
| Yes | 311 | 40.8 | 35.4 ; 46.2 | 346 | 22.2 | 17.3 ; 27.1 | 493 | 38.9 | 34.9 ; 43.0 | 53 | 21.3 | 13.4 ; 29.1 |
| No | 411 | 59.0 | 53.9 ; 64.1 | 1141 | 76.3 | 71.3 ; 81.3 | 829 | 60.7 | 56.7 ; 64.7 | 253 | 78.3 | 70.4 ; 86.1 |
| Refused | 2 | 0.2 | 0.0 ; 0.5 | 8 | 0.7 | 0.0 ; 1.5 | 3 | 0.2 | 0.0 ; 0.5 | 2 | 0.4 | 0.0 ; 1.1 |
| Don't Know | 1 | 0.1 | 0.0 ; 0.3 | 7 | 0.9 | 0.0 ; 1.7 | 1 | 0.1 | 0.0 ; 0.3 | - | - | - |
| Missing | - | - | - | - | - | - | - | - | - | 2 | 0.4 | 0.0 ; 1.1 |
| Employment Status |  |  |  |  |  |  |  |  |  |  |  |  |
| Full Time | 230 | 28.7 | 24.9 ; 32.6 | 211 | 12.2 | 8.4 ; 16.0 | 286 | 21.3 | 18.2 ; 24.3 | 33 | 13.6 | 5.5 ; 21.6 |
| Part Time | 156 | 19.5 | 14.2 ; 24.7 | 210 | 13.0 | 9.1 ; 17.0 | 341 | 23.3 | 19.7 ; 26.9 | 23 | 10.4 | 4.1 ; 16.7 |
| Not Employed | 339 | 51.8 | 43.5 ; 60.1 | 1081 | 74.8 | 69.8 ; 79.7 | 497 | 38.2 | 33.1 ; 43.3 | 143 | 40.9 | 31.8 ; 50.1 |
| Missing | - | - | - | - | - | - | 202 | 17.2 | 13.1 ; 21.3 | 109 | 35.1 | 25.0 ; 45.2 |
|  | N | % | 95% CI | N | % | 95% CI | N | % | 95% CI | N | % | 95% CI |
| Residence |  |  |  |  |  |  |  |  |  |  |  |  |
| *Rural Blantyre* | 430 | 69.2 | 51.8 ; 86.6 | 1087 | 77.8 | 64.6 ; 91.0 | 847 | 60.5 | 53.4 ; 67.7 | 223 | 66.6 | 53.9 ; 79.3 |
| *Blantyre City* | 287 | 30.0 | 12.9 ; 47.3 | 397 | 20.9 | 7.8 ; 34.0 | 443 | 37.3 | 30.2 ; 44.5 | 76 | 31.7 | 18.9 ; 44.5 |
| *Other\** | 8 | 0.7 | 0.1 ; 1.3 | 18 | 1.3 | 0.6 ; 1.9 | 36 | 2.2 | 1.0 ; 3.3 | 9 | 1.7 | 0.1 ; 3.3 |
| *Missing* | - | - | - | - | - | - | - | - | - | - | - | - |
| Sexually active in past year |  |  |  |  |  |  |  |  |  |  |  |  |
| *No* | 47 | 7.3 | 3.4 ; 11.2 | 106 | 6.9 | 3.4 ; 10.3 | 83 | 5.7 | 3.9 ; 7.5 | 18 | 6.8 | 2.7 ; 10.8 |
| *Yes* | 678 | 92.7 | 88.8 ; 96.6 | 1396 | 93.1 | 89.7 ; 96.6 | 1243 | 94.3 | 92.5 ; 96.1 | 290 | 93.2 | 89.2 ; 97.3 |
| *Missing* | - | - | - | - | - | - | - | - | - | - | - | - |
| Sex Work |  |  |  |  |  |  |  |  |  |  |  |  |
| *No* | 518 | 70.8 | 66.4 ; 75.2 | 1433 | 95.2 | 93.8 ; 96.6 | 693 | 51.3 | 47.1 ; 55.5 | 171 | 52.5 | 39.1 ; 65.9 |
| *Female sex worker* | - | - | - | 69 | 4.8 | 3.4 ; 6.2 | - | - | - | 137 | 47.5 | 34.1 ; 60.9 |
| *Male client* | 207 | 29.2 | 24.8 ; 33.6 | - | - | - | 633 | 48.7 | 44.5 ; 52.9 | - | - | - |
| *Missing* | - | - | - | - | - | - | - | - | - | - | - | - |
| Multiple Partners in Past 4 Weeks |  |  |  |  |  |  |  |  |  |  |  |  |
| *No* | 601 | 84.2 | 78.8 ; 89.6 | 1409 | 93.3 | 90.7 ; 95.9 | 935 | 69.3 | 66.2 ; 72.4 | 166 | 52.2 | 39.1 ; 65.4 |
| *Yes* | 124 | 15.8 | 10.4 ; 21.2 | 93 | 6.7 | 4.1 ; 9.3 | 391 | 30.7 | 27.6 ; 33.8 | 142 | 47.8 | 34.6 ; 60.9 |
| *Missing* | - | - | - | - | - | - | - | - | - | - | - | - |
| STI Patients |  |  |  |  |  |  |  |  |  |  |  |  |
| *No* | 605 | 81.0 | 75.0 ; 87.0 | 1264 | 83.1 | 78.7 ; 87.4 | 1136 | 86.6 | 84.4 ; 88.9 | 240 | 80.1 | 74.2 ; 86.0 |
| *Yes* | 120 | 19.0 | 13.0 ; 25.0 | 238 | 16.9 | 12.6 ; 21.3 | 190 | 13.4 | 11.1 ; 15.6 | 68 | 19.9 | 14.0 ; 25.8 |
| *Missing* | - | - | - | - | - | - | - | - | - | - | - | - |
| Sex work or multiple partners or STI patient |  |  |  |  |  |  |  |  |  |  |  |  |
| *No* | 398 | 55.3 | 49.1 ; 61.4 | 1150 | 74.9 | 69.2 ; 80.6 | 537 | 39.7 | 35.7 ; 43.7 | 130 | 42.0 | 29.5 ; 54.6 |
| *Yes* | 327 | 44.7 | 38.6 ; 50.9 | 352 | 25.1 | 19.4 ; 30.8 | 789 | 60.3 | 56.3 ; 64.3 | 178 | 58.0 | 45.4 ; 70.5 |
| *Missing* | - | - | - | - | - | - | - | - | - | - | - | - |

All number are unweighted and percentages are weighted. \*Including other regions in Malawi, Mozambique and South Africa.

### Need for prevention services and risk practices

Overall, at clinics and venues the majority of participants were sexually active in the past year (∼93%), meeting the broad criteria for needing HIV testing or condoms. This did not differ by gender. There were comparable proportions of participants reporting a STI at both sites (∼15%). Almost half (49.3%) of women at clinics were aged 15 to 24 years and 40.9% of venue participants fit the definition of adolescent girls and young women. Comparable proportions of men were aged 30 years or older (∼39%) at both sites.

As seen in Table 3 and 4, a higher proportion of participants were sex workers (47.5% vs. 4.8%) or had paid for sex with a sex worker (48.7% vs. 29.2%) in venues compared to clinics. Multiple partners were more common among women at venues (47.8%) than in the clinic (6.7%). Twice as many men from the venues reported having multiple sexual partners in the past 4 weeks compared to clinics (30.7% vs 15.8%).

**Table 3.**
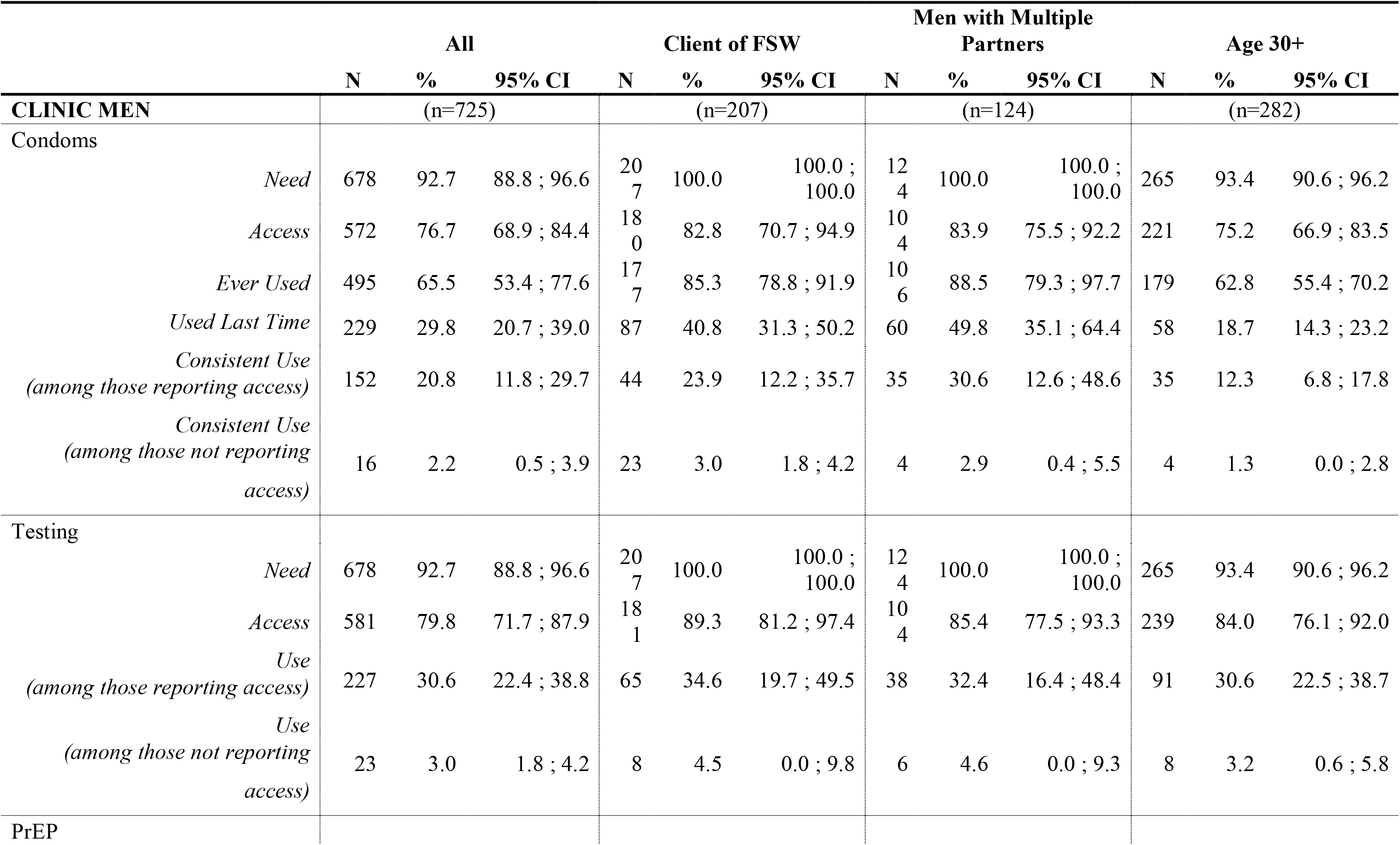
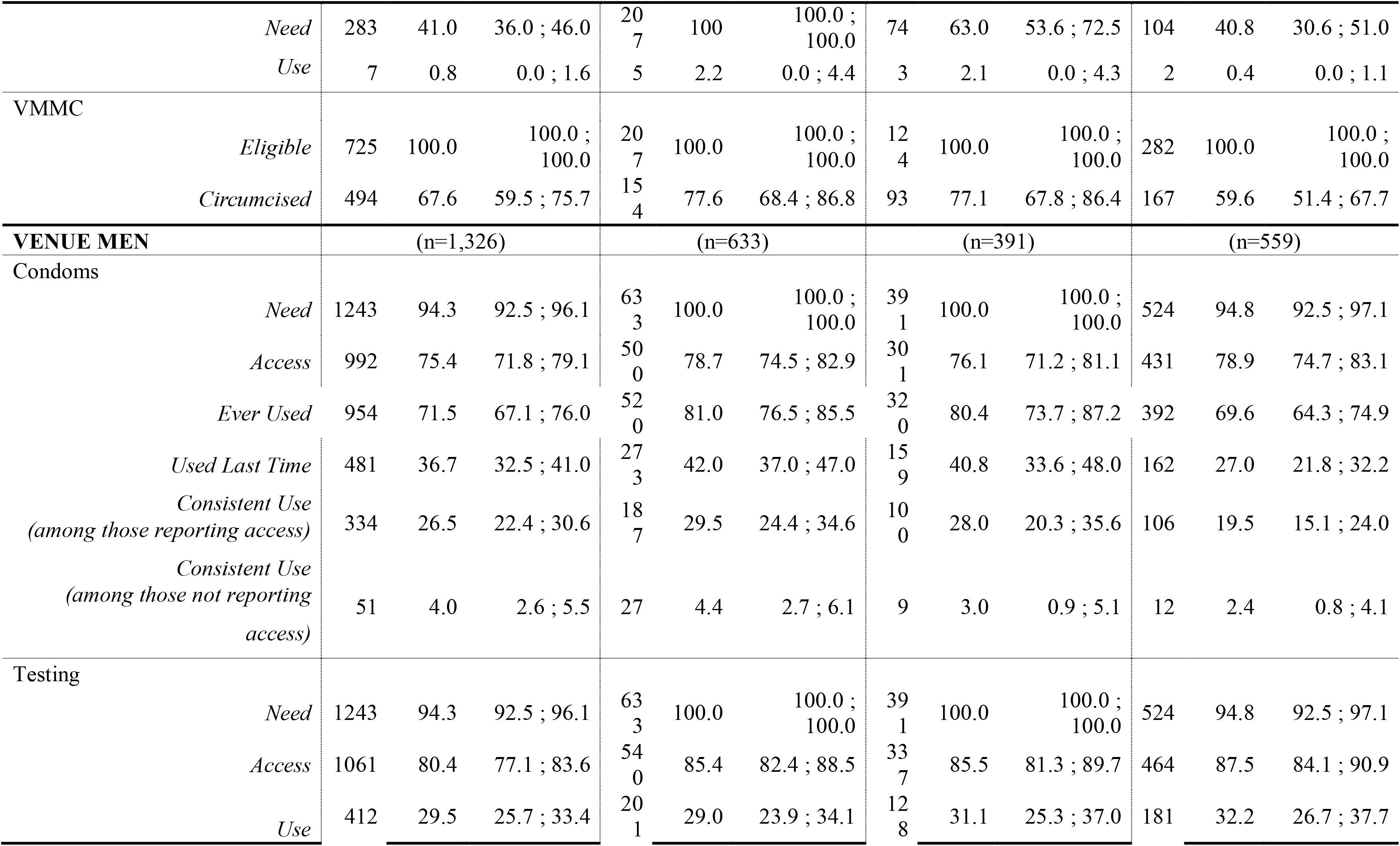
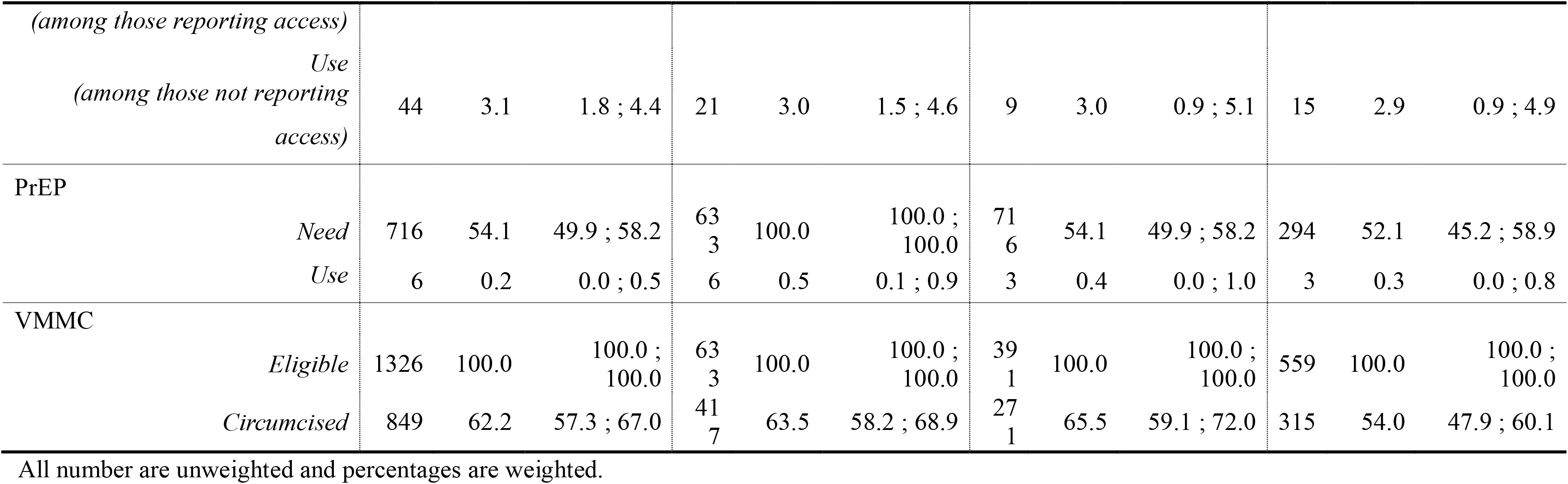
HIV Prevention Cascades among Men at Venues and at Clinics, by Risk Category (weighted %, unweighted N)

### Need, access and use of prevention services among clinic-based men by risk category

Table 3 and Fig 1 summarises need, access and use of prevention services among clinic-based men and by risk category. Among clients of FSWs, men with multiple partners and men aged 30+ reported access to condoms similarly (82.8%, 83.9% and 75.2% respectively). Use of condoms at last sex was lowest among men aged 30+ (18.7%) compared to clients of FSW and men with multiple partners (40.8% and 49.8% respectively). Consistent use, among those reporting access, ranged from 12.3% of men aged 30+ to 23.9% of male clients of FSW to 30.6% of men with multiple partners.

**Fig 1.**
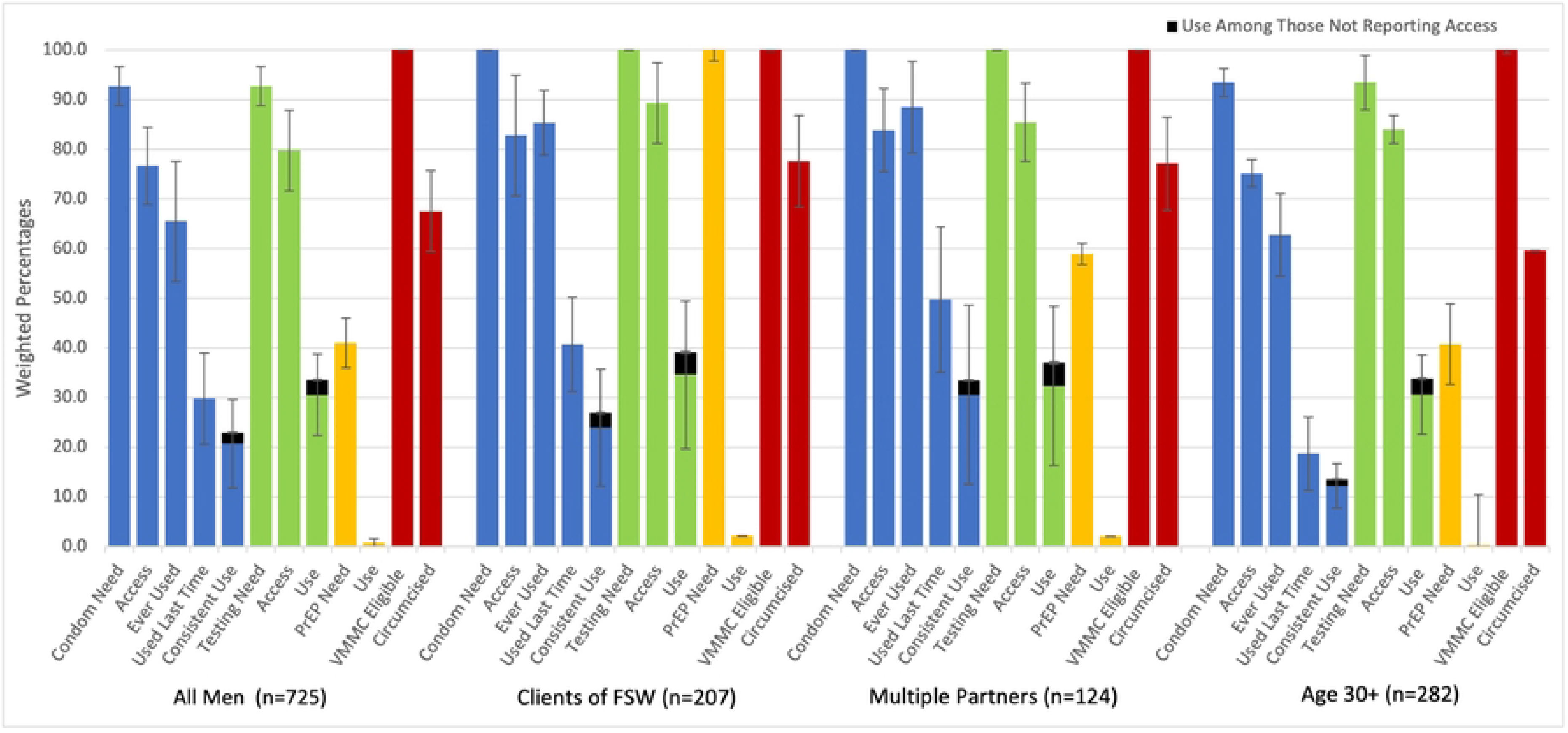
HIV Prevention Cascades for Condoms, HIV Testing, PrEP, and Voluntary Medical Male Circumcision Among Clinic Men by Risk Category. The denominator for each bar in the figures is the total number in the risk category.

Participants reported high levels of access to HIV testing across all risk groups (>80%), but with far lower levels of use (tested in the last 6 months and aware of result) of approximately 30% across all groups.

Eligibility to PrEP in this group was 41.0%, with clients of FSW all eligible (10.0%), over half of men with multiple partners eligible (63.0%) and over a third in men aged 30+ (40.8%). Among clinic-based men, 7 (0.8%) reported use with clients of FSW and men with multiple partners reporting the highest use around 2%.

Comparable proportions of men who had sex with sex workers and those with multiple partners (∼77%) reported being circumcised, but this was lower among those aged 30 years or older (59.6%).

### Need, access and use of prevention services among venue-based men by risk category

The gap between access to and use of condoms at last sex among men recruited from venues was 78.7% vs 42.0% among clients of FSW; 76.1% vs 40.8% among men with multiple partners and 78.9% vs 27.0% among men age 30+ (Table 3 and Fig 2).

**Fig 2.**
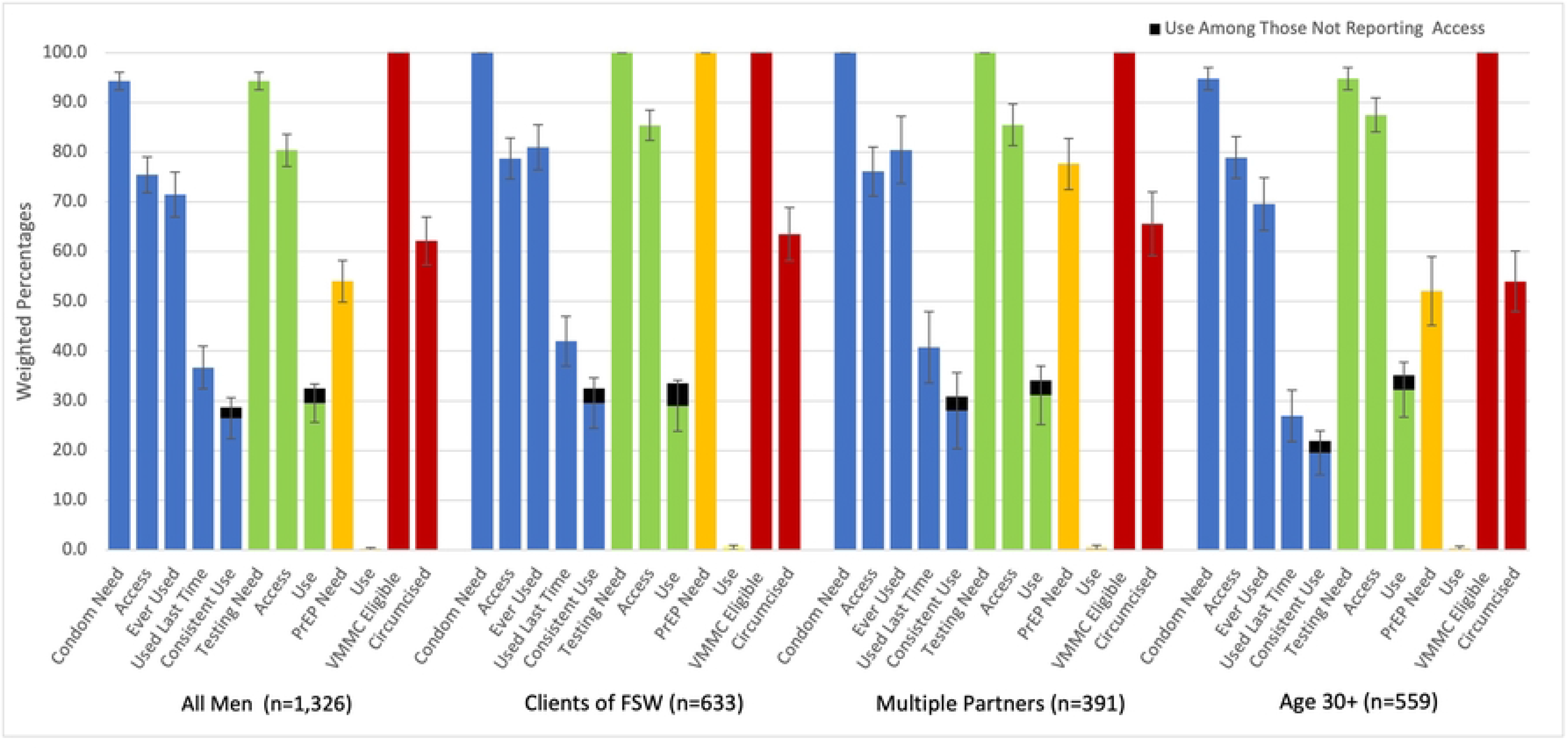
HIV Prevention Cascades for Condoms, HIV Testing, PrEP, and Voluntary Medical Male Circumcision Among Venue Men by Risk Category. The denominator for each bar in the figures is the total number in the risk category.

Access to HIV testing was high among men at venues across all risk groups (80.4%) but use among those reporting access was low (29.5%). 94.3% of venue men were eligible for PrEP with all male clients of FSW eligible (100.0%) and around half of men with multiple partners (54.1%) and men aged 30+ (52.1%) eligible. Six (0.2%) venue men reported using PrEP. Comparable proportions of male clients of FSW and those with multiple partners (∼64%) reported being circumcised, but this was lower among those aged 30 years or older (54.0%).

Overall, 23.2% of men at venues did not use any of the four prevention methods (consistent condom use, PrEP, recent HIV test or circumcision) (data not reported). The proportion ranged across risk categories from 20.1% of men who reported at least two partners in the past month to 28.6% of men aged 30 and older.

In venue-based men, the numbers reporting circumcision was lower overall compared to clinic-based men (62.2% compared to 67.6%), with those aged 30 and over at venues reporting the lowest proportion overall (54.0%).

### Need, access and use of prevention services among clinic-based women by risk category

Table 4 and Fig 3 summarises need, access and use of prevention services among clinic-based women and by risk category. 65.9% of all clinic-based women across all risk groups report that access to condoms is easy. Use of condoms at last sex was highest among FSW (43.4%) compared to women with multiple partners (38.0%) and AGYW (22.3%). Across sub-groups of women at clinics, women reporting multiple partners report the most consistent condom use among those reporting access (24.2%) compared to FSW (15.0%) and AGYW (14.5%).

**Fig 3.**
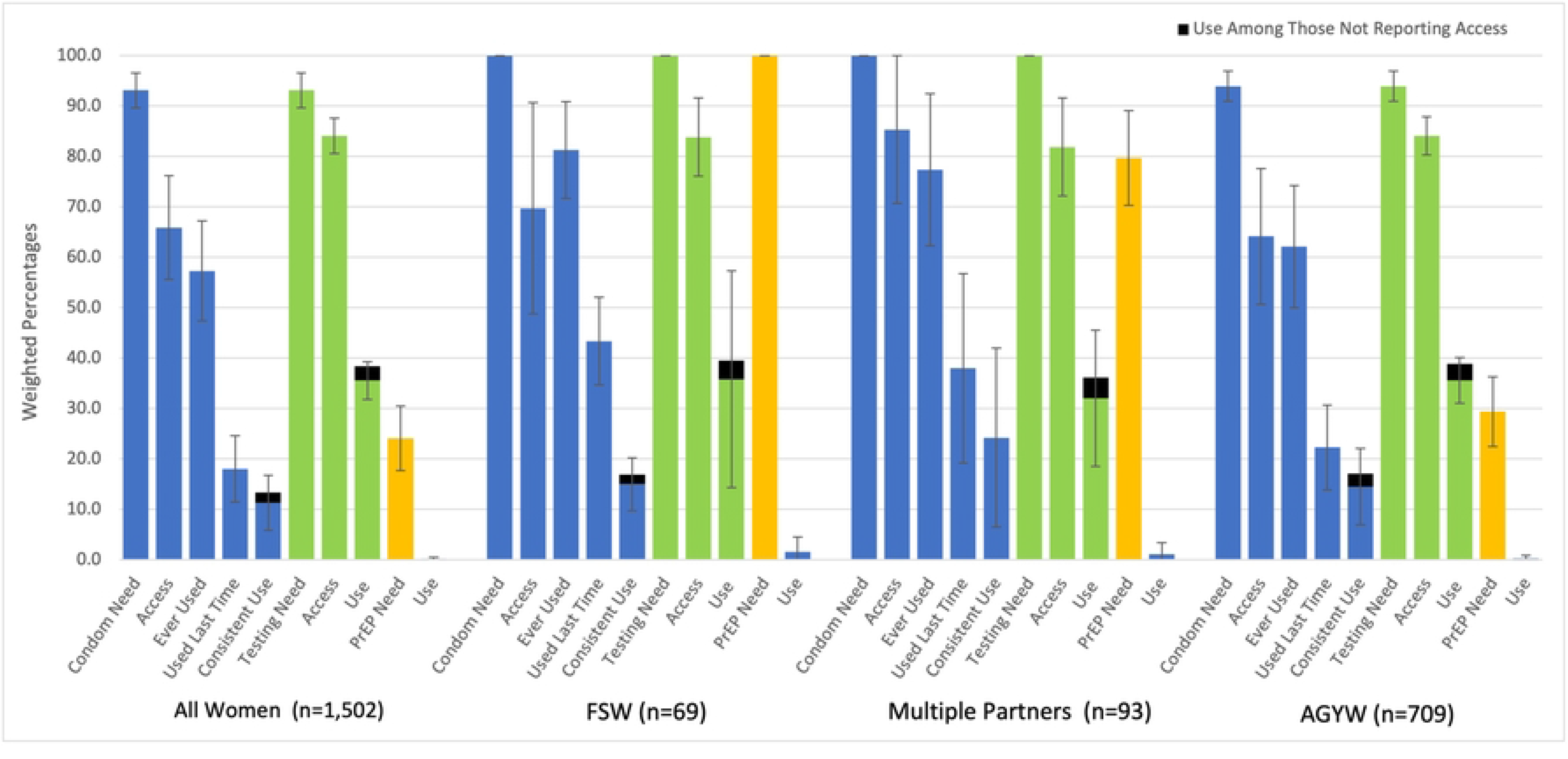
HIV Prevention Cascades for Condoms, HIV Testing, PrEP, and Voluntary Medical Male Circumcision Among Clinic Women by Risk Category. The denominator for each bar in the figures is the total number in the risk category.

**Table 4.**
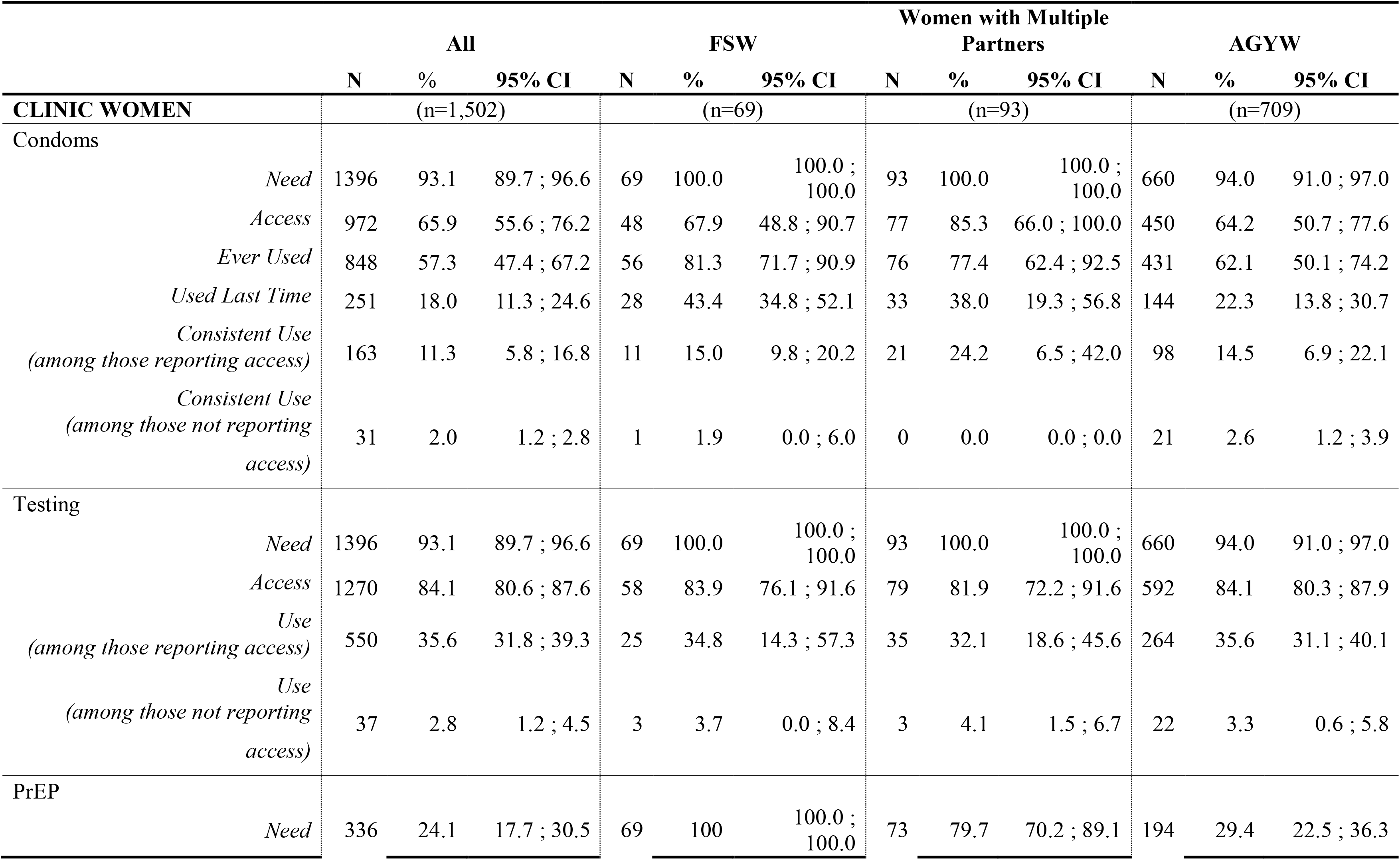
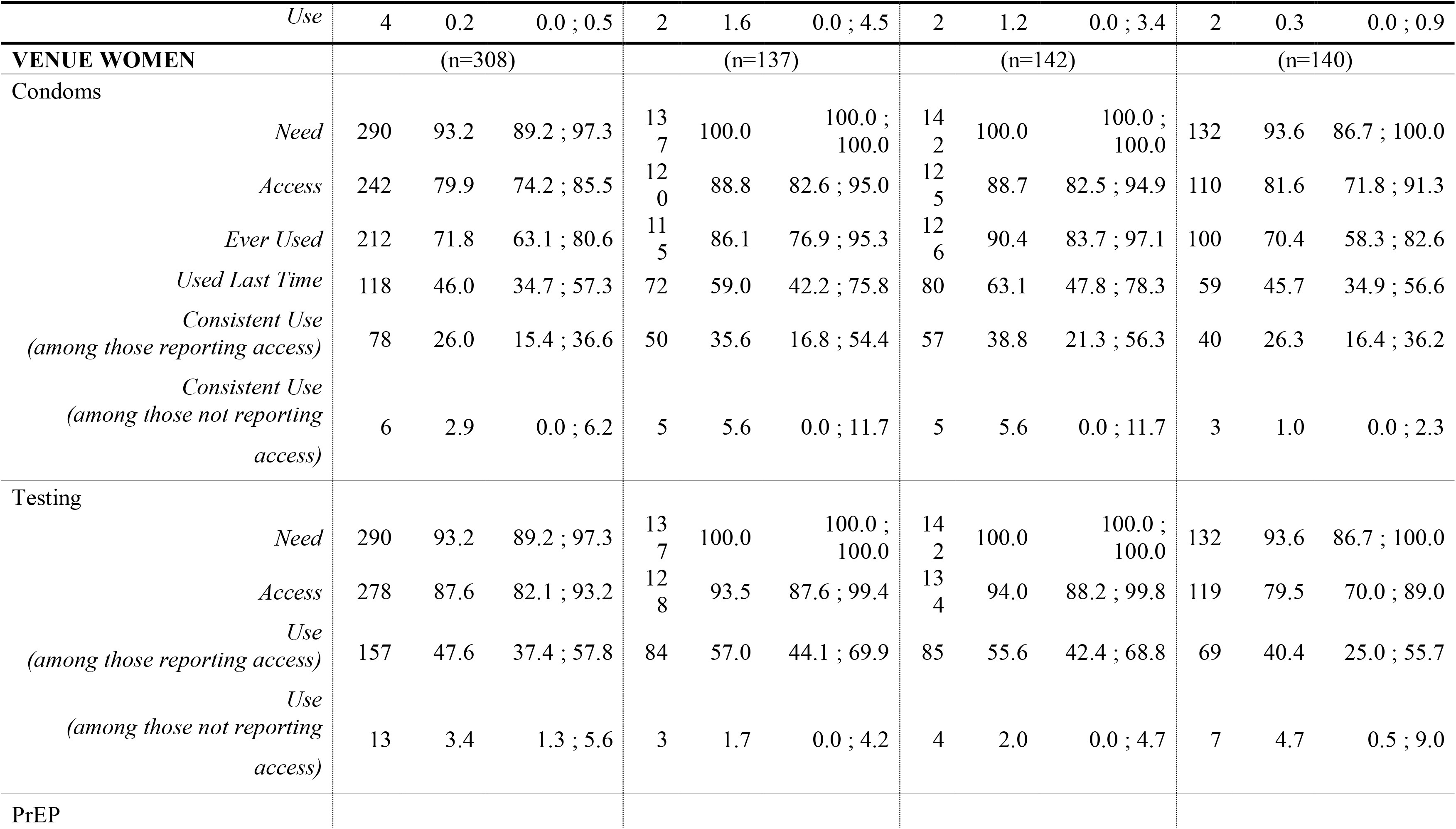
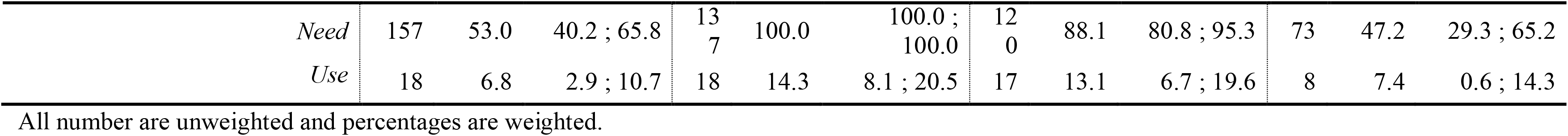
HIV Prevention Cascades among Women at Venues and at Clinics, by Risk Category (weighted %, unweighted N)

|  | All |  |  | FSW |  |  | Women with Multiple Partners |  |  | AGYW |  |  |
| --- | --- | --- | --- | --- | --- | --- | --- | --- | --- | --- | --- | --- |
|  | N | % | 95% CI | N | % | 95% CI | N | % | 95% CI | N | % | 95% CI |
| **CLINIC WOMEN** | (n=1,502) |  |  | (n=69) |  |  | (n=93) |  |  | (n=709) |  |  |
| Condoms |  |  |  |  |  |  |  |  |  |  |  |  |
| *Need* | 1396 | 93.1 | 89.7 ; 96.6 | 69 | 100.0 | 100.0 ; 100.0 | 93 | 100.0 | 100.0 ; 100.0 | 660 | 94.0 | 91.0 ; 97.0 |
| *Access* | 972 | 65.9 | 55.6 ; 76.2 | 48 | 67.9 | 48.8 ; 90.7 | 77 | 85.3 | 66.0 ; 100.0 | 450 | 64.2 | 50.7 ; 77.6 |
| *Ever Used* | 848 | 57.3 | 47.4 ; 67.2 | 56 | 81.3 | 71.7 ; 90.9 | 76 | 77.4 | 62.4 ; 92.5 | 431 | 62.1 | 50.1 ; 74.2 |
| *Used Last Time* | 251 | 18.0 | 11.3 ; 24.6 | 28 | 43.4 | 34.8 ; 52.1 | 33 | 38.0 | 19.3 ; 56.8 | 144 | 22.3 | 13.8 ; 30.7 |
| *Consistent Use*<br>*(among those reporting access)* | 163 | 11.3 | 5.8 ; 16.8 | 11 | 15.0 | 9.8 ; 20.2 | 21 | 24.2 | 6.5 ; 42.0 | 98 | 14.5 | 6.9 ; 22.1 |
| *Consistent Use*<br>*(among those not reporting access)* | 31 | 2.0 | 1.2 ; 2.8 | 1 | 1.9 | 0.0 ; 6.0 | 0 | 0.0 | 0.0 ; 0.0 | 21 | 2.6 | 1.2 ; 3.9 |
| Testing |  |  |  |  |  |  |  |  |  |  |  |  |
| *Need* | 1396 | 93.1 | 89.7 ; 96.6 | 69 | 100.0 | 100.0 ; 100.0 | 93 | 100.0 | 100.0 ; 100.0 | 660 | 94.0 | 91.0 ; 97.0 |
| *Access* | 1270 | 84.1 | 80.6 ; 87.6 | 58 | 83.9 | 76.1 ; 91.6 | 79 | 81.9 | 72.2 ; 91.6 | 592 | 84.1 | 80.3 ; 87.9 |
| *Use*<br>*(among those reporting access)* | 550 | 35.6 | 31.8 ; 39.3 | 25 | 34.8 | 14.3 ; 57.3 | 35 | 32.1 | 18.6 ; 45.6 | 264 | 35.6 | 31.1 ; 40.1 |
| *Use*<br>*(among those not reporting access)* | 37 | 2.8 | 1.2 ; 4.5 | 3 | 3.7 | 0.0 ; 8.4 | 3 | 4.1 | 1.5 ; 6.7 | 22 | 3.3 | 0.6 ; 5.8 |
| PrEP |  |  |  |  |  |  |  |  |  |  |  |  |
| *Need* | 336 | 24.1 | 17.7 ; 30.5 | 69 | 100 | 100.0 ; 100.0 | 73 | 79.7 | 70.2 ; 89.1 | 194 | 29.4 | 22.5 ; 36.3 |
|  | N | % | 95% CI | N | % | 95% CI | N | % | 95% CI | N | % | 95% CI |
| *Use* | 4 | 0.2 | 0.0 ; 0.5 | 2 | 1.6 | 0.0 ; 4.5 | 2 | 1.2 | 0.0 ; 3.4 | 2 | 0.3 | 0.0 ; 0.9 |
| **VENUE WOMEN** | (n=308) |  |  | (n=137) |  |  | (n=142) |  |  | (n=140) |  |  |
| Condoms |  |  |  |  |  |  |  |  |  |  |  |  |
| *Need* | 290 | 93.2 | 89.2 ; 97.3 | 137 | 100.0 | 100.0 ; 100.0 | 142 | 100.0 | 100.0 ; 100.0 | 132 | 93.6 | 86.7 ; 100.0 |
| *Access* | 242 | 79.9 | 74.2 ; 85.5 | 120 | 88.8 | 82.6 ; 95.0 | 125 | 88.7 | 82.5 ; 94.9 | 110 | 81.6 | 71.8 ; 91.3 |
| *Ever Used* | 212 | 71.8 | 63.1 ; 80.6 | 115 | 86.1 | 76.9 ; 95.3 | 126 | 90.4 | 83.7 ; 97.1 | 100 | 70.4 | 58.3 ; 82.6 |
| *Used Last Time* | 118 | 46.0 | 34.7 ; 57.3 | 72 | 59.0 | 42.2 ; 75.8 | 80 | 63.1 | 47.8 ; 78.3 | 59 | 45.7 | 34.9 ; 56.6 |
| *Consistent Use*<br>*(among those reporting access)* | 78 | 26.0 | 15.4 ; 36.6 | 50 | 35.6 | 16.8 ; 54.4 | 57 | 38.8 | 21.3 ; 56.3 | 40 | 26.3 | 16.4 ; 36.2 |
| *Consistent Use*<br>*(among those not reporting access)* | 6 | 2.9 | 0.0 ; 6.2 | 5 | 5.6 | 0.0 ; 11.7 | 5 | 5.6 | 0.0 ; 11.7 | 3 | 1.0 | 0.0 ; 2.3 |
| Testing |  |  |  |  |  |  |  |  |  |  |  |  |
| *Need* | 290 | 93.2 | 89.2 ; 97.3 | 137 | 100.0 | 100.0 ; 100.0 | 142 | 100.0 | 100.0 ; 100.0 | 132 | 93.6 | 86.7 ; 100.0 |
| *Access* | 278 | 87.6 | 82.1 ; 93.2 | 128 | 93.5 | 87.6 ; 99.4 | 134 | 94.0 | 88.2 ; 99.8 | 119 | 79.5 | 70.0 ; 89.0 |
| *Use*<br>*(among those reporting access)* | 157 | 47.6 | 37.4 ; 57.8 | 84 | 57.0 | 44.1 ; 69.9 | 85 | 55.6 | 42.4 ; 68.8 | 69 | 40.4 | 25.0 ; 55.7 |
| *Use*<br>*(among those not reporting access)* | 13 | 3.4 | 1.3 ; 5.6 | 3 | 1.7 | 0.0 ; 4.2 | 4 | 2.0 | 0.0 ; 4.7 | 7 | 4.7 | 0.5 ; 9.0 |
| PrEP |  |  |  |  |  |  |  |  |  |  |  |  |

|  |  | All |  |  | FSW |  |  | Women with Multiple Partners |  |  | AGYW |  |  |
| --- | --- | --- | --- | --- | --- | --- | --- | --- | --- | --- | --- | --- | --- |
|  |  | N | % | 95% CI | N | % | 95% CI | N | % | 95% CI | N | % | 95% CI |
|  | *Need* | 157 | 53.0 | 40.2 ; 65.8 | 137 | 100.0 | 100.0 ; 100.0 | 120 | 88.1 | 80.8 ; 95.3 | 73 | 47.2 | 29.3 ; 65.2 |
|  | *Use* | 18 | 6.8 | 2.9 ; 10.7 | 18 | 14.3 | 8.1 ; 20.5 | 17 | 13.1 | 6.7 ; 19.6 | 8 | 7.4 | 0.6 ; 14.3 |

All number are unweighted and percentages are weighted.

Overall, a high proportion of clinic-based women report easy access to HIV testing (84.1%), but lower proportions have been tested in the last 6 months among those reporting access (35.6%). In our sample, 24.1% of clinic-based women would qualify for PrEP services based on the national guidelines (Table 1) but uptake is low (0.2%). Only two clinic-based women (n=69) who reported sex work were on PrEP, although sex workers are a priority group for PrEP.

### Need, access and use of prevention services among venue-based women by risk category

Easy access to condoms was reported by 79.9% of all venue-based women as shown in Table 4 and Fig 4. Use of condoms at last sex is at 46.0% and varies by risk group: 59.0% among clinic women who report sex work, 63.1% among women with multiple partners and 45.7% among AGYW. Approximately a third of FSW (35.6%), women with multiple partners (38.8%) and AGYW (26.3%) reported consistent condom use among those reporting access.

**Fig 4.**
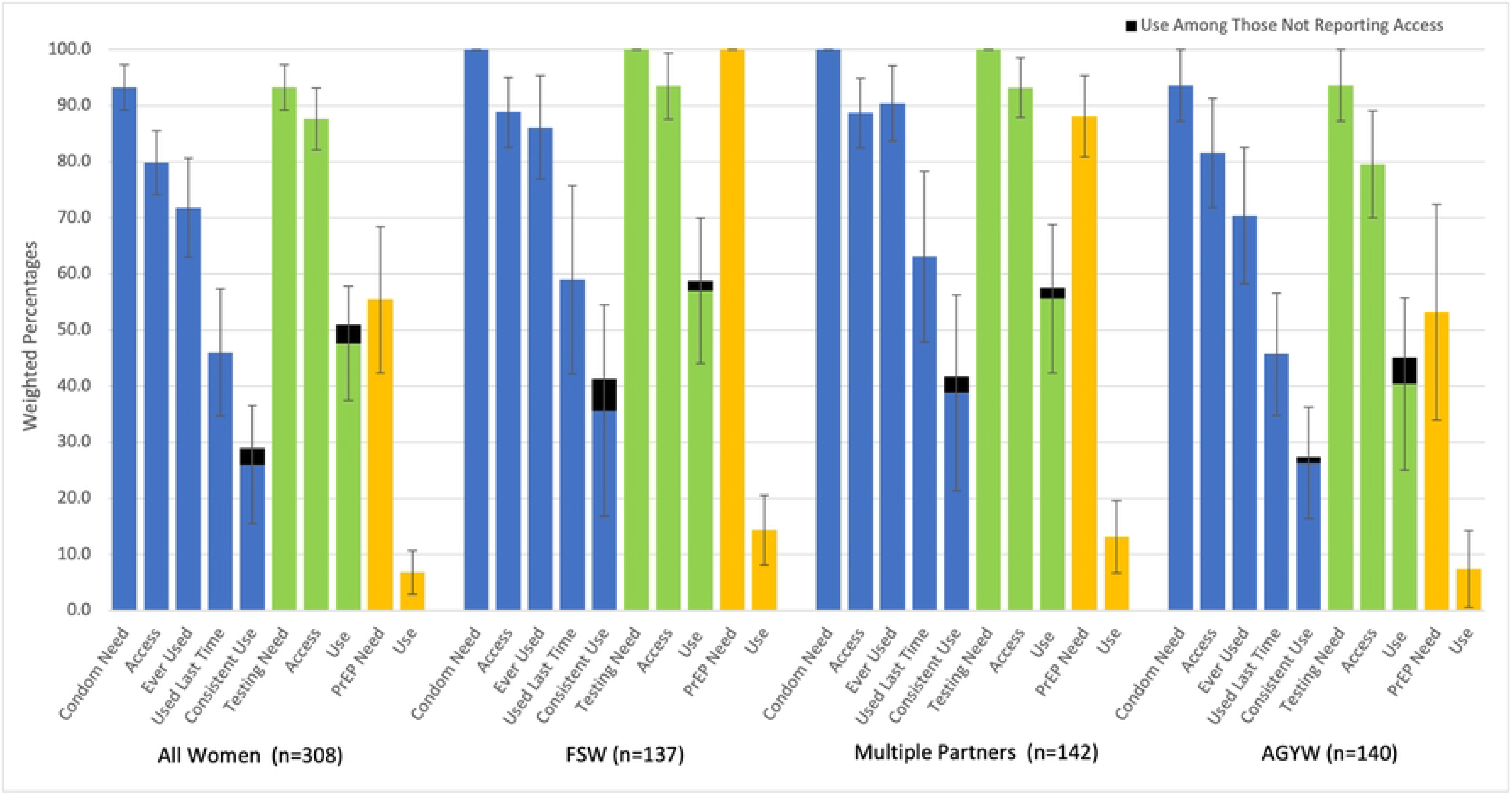
HIV Prevention Cascades for Condoms, HIV Testing, PrEP, and Voluntary Medical Male Circumcision Among Venue Women by Risk Category. The denominator for each bar in the figures is the total number in the risk category.

Access to HIV testing is high across all risk groups (87.6%), and lowest among AGYW (79.5%) compared to FSW (93.5%) and women with multiple partners (94.0%). Around half (47.6%) of venue women reported testing for HIV in the last 6 months and this was lowest among AGYW (40.4%) compared to FSW (57.0%) and women with multiple partners (55.6%). Overall just over half (53.0%) of women at venues qualify for PrEP services and use of PrEP is higher (6.8%) compared to clinic women. Reported PrEP use is highest among FSW (14.3%) and women with multiple partners (13.1%).

## Discussion

We found marked prevention service gaps for condoms, HIV testing, PrEP and circumcision for participants at both clinics and venues. A key contribution of this study is the direct comparison of prevention gaps across the two settings. Venue participants reported substantially higher risk practices in relation to sex work (among women), paying for sex (among men) and having multiple partners (among both). This argues for prioritising prevention at venues. However, clinics also serve large numbers of AGYW and individuals with unmet prevention needs who may not be reached through venue-based approaches alone. Though reported access to condoms and HIV testing was high, uptake of these services was considerably lower, indicating barriers to uptake or lack of motivation. The most striking gap was for PrEP. Uptake was approximately 2% overall and remained low even among priority groups such as venue-based female sex workers (14.3%) and male clients of sex workers (2.2% at clinics and 0.5% at venues). These findings are consistent with the early stage of PrEP roll-out in Malawi at the time of data collection (late 2021) and provide an important baseline against which future scale-up should be measured. Beyond PrEP, gaps were also pronounced among AGYW and older men across multiple prevention services.

We identified gaps in access in the condom, HIV testing and PrEP cascades. Around a quarter of men and women at clinics and venues reported that access to condoms was not easy. A fifth of people at clinics reported not knowing where to get an HIV test in Blantyre even though they were being seen in a clinic that provided testing and routinely offered condoms in testing and STI services. Our findings document that half of venue-based women and almost half of clinic and venue-based men are eligible for PrEP based on STI history and involvement with sex work. PrEP delivery in Malawi is at the early stages, included in the National HIV Prevention Strategy 2015-2020 but not promoted significantly until late 2021 [22]. Consequently only 2% of participants were on PrEP at the time of the study limiting our ability to assess access and uptake accurately. Other research in the region has found high interest in PrEP [23–29]. In terms of circumcision, uptake was around 60%, but lowest among older men in both clinics and venues. Perceived access to circumcision was not assessed in this study but the circumcision levels we found were much higher than the 2015-2016 Malawi Demographic and Health Survey figure of 28% circumcision for men 15-49 [30]. A 2021 study in Machinga district found that some barriers to uptake of VMMC were age, accessibility, cultural influences and fear of complications such as pain and bleeding [31]. Indeed, we also found circumcision levels to be consistently lower in older men across both venue and clinic participants. A focus on increasing uptake of circumcision in older men through understanding barriers to access and uptake would be an important next step.

In the condom cascade, even though access to condoms was generally high, consistent condom use (among those reporting access) was generally low, with venue-recruited female sex workers and women with multiple partners reporting the highest proportion of consistent condom use around 35%. Comparable low levels of condom use have been reported elsewhere in the region, despite good access [14,16]. In the 2024 Malawi DHS, 45.2% of women who reported higher-risk sex (with a non-marital or non-cohabiting partner) used a condom at last sex [32]. Among men who had paid for sex, we found much lower use of condom at last sex (40.8% in clinics and 42.0% in venues) compared to what was reported in the 2015-16 Malawi DHS (75.1%)[30]. These comparisons should be interpreted with caution given differences in the populations and condom-use measures compared and the sampling methodologies. Among men and women with multiple partners, we found a higher use of condoms at last sex, (49.8% in clinic men and 40.8% in venue men; 38.0% in clinic women and 63.1% in venue women), compared to the 2024 Malawi DHS (38.4% in men and 30.6% in women) [32]. Such findings highlight a positive change in condom use among men and women with multiple partners, but a decrease among men paying for sex indicating that though these two groups often overlap, there may be different factors that affect choice of condom use. The role of structural factors hindering condom use, such as unequal financial, gender and power dynamics, poverty, alcohol use and stigma or policing practices around transactional sex have been well documented and should be addressed to create environments in which condom use is accepted [16,33–36]. Although access should not be discounted as a factor in condom use, our study shows that, those reporting access to condoms reported higher consistent use of condoms than those without easy access. In Blantyre, there are challenges at every step in the condom cascade—access, first use, and consistent use. Even with increasing access to treatment, condoms remain a critical prevention option for protection against STIs and pregnancy prevention, particularly in the context of high prevalence of syphilis, chlamydia and gonorrhoea among sex workers [37].

Our findings suggest lower uptake of HIV testing among venue-based participants (34.0%) and clinic attenders (34.3%) than levels reported in the latest population-based HIV impact assessment (45.2% of adults aged 15 years and older) [2]. However, women at venues (& reporting access) reported the highest uptake of HIV testing at 47.6%, which is above the national average. Other studies have reported varied uptake of HIV testing in Malawi [2,13,38]. A 2018 venue-based study in Zomba reported 75% of women engaging in transactional sex had been tested for HIV in the last 6 months, compared to 59% of venue-based FSW in our study [13]. In Malawi, HIV testing services are provided through healthcare facilities, community-based programs, and outreach efforts. As with condoms, access was associated with use. Those who knew where to get tested had reported being tested in higher numbers compared to those who did not know where to get tested. Understanding why all individuals, but specifically men and women at clinics, lack access, ability or motivation to take up HIV testing is crucial in sustainably expanding provision of HIV testing to venue participants [13].

There is a wealth of research documenting HIV prevention cascades since the cascade was first conceptualised, and the operationalization of the cascades has varied in relation to prevention method, measurement of motivation, access and use, and reference population, making comparisons difficult [39]. It is also important to note that the definition of female sex work used in this study captured any exchange of sex for gifts, favours or money in the past 12 months. This broad definition includes informal and occasional transactional sex alongside more regular sex work [40–42]. Women for whom sex work is a primary income source likely face higher HIV risk and may have different prevention needs than those who engage in occasional transactions [43]. Future studies should consider stratifying by frequency of transactional sex and number of partners to distinguish these groups more precisely. In contrast to other studies, we did not measure motivation for prevention, further limiting comparisons with other evidence. We were not able to collect data on access to PrEP and circumcision and may have also over-simplified the measure of access for HIV testing and condoms compared to other prevention cascade papers in the region [44–46]. The time-location sampling design captures individuals present at mapped venues and clinics during defined survey periods. Our findings should therefore be interpreted within this sampling frame and not generalised to the broader population of Blantyre or to individuals who do not frequent these settings. The survey recruitment approach may also have oversampled daytime venue patrons relative to those present at night. This could underestimate risk behaviours among the most active sex workers who predominantly operate during evening and nighttime hours [21]. Additionally, comparisons with the 2015-16 Malawi DHS should be interpreted with caution given differences in sampling methodology and the eight-year interval between surveys. The strengths of our study lie in the use of the PLACE protocol, which has been implemented over time and in different countries across the region making data easier to compare as questions and methodologies are consistent [13,47]. Additionally, we had a very large population across both clinics and venues. Going to venues to carry out the survey meant that we were able to reach populations at venues who may not often or ever access the clinic.

## Conclusions

This is one of the first studies in Malawi to simultaneously compare HIV prevention service gaps across multiple populations, prevention modalities and service delivery settings. Gaps in prevention uptake persist among individuals reachable at both public health facilities and social venues. The most critical gap is for PrEP. Uptake was suboptimal across almost all groups despite high eligibility, and this finding should inform the ongoing scale-up of PrEP delivery in Malawi. Among other prevention services, AGYW at venues and older men at both settings reported consistently lower use than individuals engaged in sex work or those with multiple partners. Clinic-based services should be tailored to better engage AGYW, given their high eligibility and low uptake of PrEP and HIV testing in this setting. New approaches that involve key and vulnerable population leaders and social venue owners to link communities to clinics and national programmes may help bridge these gaps

## Competing interests

The authors declare no competing interest. The study reported in this publication was supported by the MeSH Consortium which is funded by the Bill & Melinda Gates Foundation. The content is solely the responsibility of the authors and does not necessarily represent the official views of funders.

## Authors’ contributions

SSW, MCH, JRH, BR and LP conceived the study. SSW, ES, JC, CB and EK collected and prepared the data for analysis. CB and SB led the analysis of the data, with analysis contributions by SSW, CB, ES, JRH, TH, HJ, and LP. CB, SB, ES and SSW led the interpretation and manuscript write-up, with contributions from all authors. All authors reviewed the manuscript, provided inputs, and approved the final manuscript.

## Acknowledgements

We acknowledge the important work done by our 35 Research Assistants during three months of data collection and HIV testing in Blantyre, largely during rainy season. Also, the CLOVE study was implemented in collaboration with the Blantyre Prevention Strategy (BPS) which was launched in 2020 by the Malawi Ministry of Health in collaboration with Georgetown University, working closely with all PEPFAR-funded implementation partners in Blantyre. The BPS goals are to support establishment of a district-based system that will enhance deployment and uptake of novel and existing HIV prevention interventions and products and to have HIV prevention institutionalized as a cohesive, effective, and sustainable country-led response, with coordinated external support.

## Funding

The study was carried out by the MeSH Consortium at UNC Project Malawi and London School of Hygiene & Tropical Medicine-UK, which is funded by the Bill & Melinda Gates Foundation grant number INV-007055. The content is solely the responsibility of the authors and does not necessarily represent the official views of funders.

## Data Availability Statement

The data that support the findings of this study are available on request from the corresponding author. The data are not publicly available due to privacy or ethical restrictions.

## Supporting Information

**S1 File. Information on the CLOVE study and methodology.**

**S1 Fig 1. CLOVE study clinic selection process**

**S1 Fig 2. CLOVE Study venue sampling design**

## Notes

### Competing Interest Statement

The authors have declared no competing interest.

